# Longitudinal trajectories of cortical folding in schizophrenia spectrum disorders: a 13-year follow-up study

**DOI:** 10.1101/2025.06.10.25329046

**Authors:** Lara A. Wallenwein, Laura A. Wortinger, Claudia Barth, Pravesh Parekh, Kjetil N. Jørgensen, Lena Palaniyappan, Daniela Mier, Erik G. Jönsson, Ingrid Agartz, Stener Nerland

**Author notes:** Corresponding author: Stener Nerland.

## Abstract

**Background and Hypothesis:** Altered cortical folding is a putative marker of neurodevelopmental disruption in schizophrenia spectrum disorders (SSD). Patients with SSD have been hypothesized to exhibit an accelerated decline in age-related cortical folding, quantified with the Local Gyrification Index (LGI). Here, we assessed longitudinal and cross-sectional LGI differences in patients with chronic SSD relative to healthy controls across 13 years.

**Study Design:** The sample comprised patients with SSD (mean baseline age=41.28 years) and healthy controls (mean baseline age=41.56 years) with MRI acquisitions at baseline (103 SSD; 99 controls) and follow-up after 5 (50 SSD; 57 controls) and 13 years (42 SSD; 60 controls). T1-weighted images were processed with the longitudinal pipeline in FreeSurfer. Spatio-temporal linear mixed-effects models were used to test for longitudinal and cross-sectional case-control differences in LGI, as well as the impact of symptom severity and antipsychotic medication dose among patients.

**Study Results:** Although cross-sectional LGI was lower in patients in extensive frontal, parietal, and occipital regions, we observed no significant differences in longitudinal trajectories between patients and controls after FDR correction. Medication dose was linked cross-sectionally to lower LGI of the anterior cingulate and orbitofrontal cortex, and the postcentral gyrus.

**Conclusion:** In the longest longitudinal study on cortical folding in SSD to date, we found no accelerated progressive decline in cortical folding in patients. Thus, chronic SSD appears to be characterized by a state of stable hypogyria relative to healthy controls, supporting the interpretation of LGI as a marker of neurodevelopmental or early-life disturbances in SSD.

## Introduction

Altered cortical brain morphology is a well-established feature of schizophrenia spectrum disorders (SSD), believed to stem from neurodevelopmental disruptions.^1–3^ Gyrification, i.e., the process that gives rise to the characteristic sulci and gyri of the cerebral cortex, is frequently quantified using the Local Gyrification Index (LGI).^4^ Aberrant gyrification has been proposed as a marker of neurodevelopmental disruption in SSD, since it is mainly determined during fetal development.^5,6^ However, cortical folding changes have also been reported after birth and therefore reflect both static and dynamic components.^7–9^ In healthy individuals, cortical folding has been shown to decline with age during mid-to-late adulthood, reflecting normal age-related changes.^10–12^

Numerous studies have reported altered cortical folding in SSD, though the direction of these effects varies by illness stage,^13^ potentially indicating an illness-related neurodegenerative decline.^14^ It has been argued that at-risk individuals and patients in the early stages of illness exhibit higher cortical folding (hypergyria), while chronic SSD is associated with lower cortical folding (hypogyria) compared to healthy controls.^2,13,15^ Supporting this notion, some studies have reported hypergyria in early illness stages,^16,17^ and cross-sectional studies of adults with SSD have observed hypogyria.^18^ To reconcile these findings, it has been hypothesized that SSD involves an accelerated decline in cortical folding beyond normal age-related reductions.^10,13,15^ However, the cross-sectional design of most prior studies precludes the assessment of progressive illness-related changes, underscoring the need for longitudinal investigations of gyrification in this population.

To date, there have been few longitudinal studies on cortical folding in SSD. A study on patients with first-episode psychosis found greater LGI reductions among patients compared to healthy controls in frontal, temporal, and parietal areas over 2.7 years.^19^ A second study reported greater LGI decreases in the left insula and inferior frontal cortex in adolescent-onset psychosis compared to healthy controls over 2 years.^20^ However, another study found no longitudinal group differences in global LGI after a 6-week medication trial.^21^ Other longitudinal studies on cortical folding did not directly compare patients and healthy controls or employed network measures.^17,22,23^

Antipsychotic medication and symptom severity may impact the trajectory of brain morphology changes. However, prior findings have been inconsistent, with reports of negative, ^24^ positive^17^ or no cross-sectional associations between cortical folding and medication.^25–27^ and no associations with longitudinal folding trajectories.^19^ Similarly, cross-sectional associations between cortical folding and psychopathology have yielded mixed results.^24,26–30^ Longitudinal gyrification change has also been linked to symptom severity, although with inconsistent directions and location of effects.^17,19,21^ Thus, further research on longitudinal associations with antipsychotic medication and psychotic symptom severity in well-powered datasets is warranted.

Here, we investigated both longitudinal and cross-sectional changes in cortical folding, measured with LGI, in a sample of chronic patients with SSD and healthy controls, with follow-up assessments conducted 5 and 13 years after baseline. Based on prior literature, we expected greater LGI decrease among patients with SSD compared to controls. Within the patient group, we further assessed longitudinal and cross-sectional associations with symptom severity and antipsychotic medication dose. Given the mixed findings in previous studies, these analyses were exploratory in nature. To ensure the robustness of our results, we performed extensive validations with respect to processing method, statistical modelling, and data quality.

## Materials and methods

### Participants

Participants were recruited between 1999 and 2003 as part of the Human Brain Informatics (HUBIN) project at Karolinska Institutet in Stockholm, Sweden. The HUBIN study was approved by the Swedish Ethical Review Authority (Etikprövningsmyndigheten) and the Research Ethics Committee at Karolinska Institutet. Data handling procedures were approved by the Norwegian Data Protection Authority (Datatilsynet; #2003/2052) and the Data Protection Officers (PVO) of the Oslo University Hospital. The study was conducted in compliance with the General Data Protection Regulation (GDPR) and the Declaration of Helsinki. Patients with SSD were referred from catchment areas within the North-Western Stockholm County and healthy controls were selected from the Swedish Population Register and recruited among hospital staff. Inclusion criteria required participants to be aged 18-65 years at baseline with an IQ above 70. Exclusion criteria included neurological illness or a history of moderate-to-severe head injury, and for healthy controls a history of substance abuse or dependency or having a first-degree relative with a severe mental disorder.

Clinical assessment was performed using the SCID-III-R,^31^ Schedules for Clinical Assessment in Neuropsychiatry (SCAN),^32^ the Scales for the Assessment of Positive and Negative Symptoms (SAPS and SANS),^33,34^ and reviews of medical records.^35–37^ Antipsychotic medication use was determined in interviews and converted to chlorpromazine-equivalent doses (CPZ).^38^ Age at onset was defined as age of first psychotic episode and duration of illness as years from age at onset to age at Magnetic Resonance Imaging (MRI) examination. IQ was estimated with the Wechsler Adult Intelligence Scale-III (WAIS-III) Vocabulary subtest.^39,40^ Alcohol and illicit drug use were assessed in interviews and using the Alcohol Use Disorder Identification Test (AUDIT).^41,42^ The Global Assessment of Functioning Scale (GAF)^43^ was used to assess psychosocial functioning. The clinical protocols were repeated at each follow-up time point. Clinical assessments at each time point were conducted by the same psychiatrist (E.G.J.).

The final study sample included 103 patients with SSD and 99 healthy controls at baseline, 50 patients and 57 healthy controls at the 5-year follow-up, and 42 patients and 60 healthy controls at 13-year follow-up (Table 1). Out of the 103 patients included in the analyses, 76 patients were diagnosed with schizophrenia (73.8%), 16 with schizoaffective disorder (15.5%), and 11 with other psychotic disorders (10.7%).

**Table 1.** Sample characteristics for each time point.

|  | Patients with SSD |  |  | Healthy controls |  |  |
| --- | --- | --- | --- | --- | --- | --- |
|  | Baseline | 5-year follow-up | 13-year follow-up | Baseline | 5-year follow-up | 13-year follow-up |
| n | 103 | 50 | 42 | 99 | 57 | 60 |
| % female | 28.2% | 20.0% | 23.8% | 35.4% | 35.1% | 38.3% |
| Age (years) | 41.28 (7.68) | 45.63 (7.37) | 51.46 (8.1) | 41.56 (9.06) | 47.61 (7.89) | 55.47 (8.34) |
| Duration to follow-up (years) | - | 5.17 (0.43) | 13.14 (0.73) | - | 5.17 (0.23) | 13.13 (0.49) |
| % right-handed | 86.0% | 91.8% | 85.0% | 92.8% | 87.5% | 88.1% |
| Years of schooling | 13.4 (9.38) | 12.5 (2.35) | 12.71 (2.37) | 14.31 (2.96) | 14.23 (2.45) | 14.69 (3.06) |
| IQ at baseline | 89.54 (19.47) | 92.01 (18.4) | 93.22 (15.74) | 103.75 (14.38) | 104.14 (14.42) | 103.09 (14.25) |
| Duration of illness (years) | 16.37 (8.96) | 21.5 (8.35) | 27.52 (8.43) | - | - | - |
| Age at onset (years) | 24.77 (6) | 23.79 (4.24) | 23.52 (4.63) | - | - | - |
| GAF | 50.71 (10.8) | 47.71 (11.15) | 44.63 (11.48) | 87.45 (5.17) | 84.45 (8.95) | 81.95 (10.09) |
| SAPS | 10.21 (9.83) | 9.32 (9.2) | 9.83 (8.44) | - | - | - |
| SANS | 21.18 (14.26) | 24.96 (16.41) | 24.41 (14.8) | - | - | - |
| CPZ | 263.58 (257.58) | 255.82 (213.76) | 328.05 (283.85) | - | - | - |

*Note.* Mean and standard deviations reported for continuous variables and percentages for categorical variables. Summary metrics were calculated for non-missing data.
*Abbreviations:* SSD = Schizophrenia Spectrum Disorder; GAF = Global Assessment of Functioning scale; SAPS = Scale for the Assessment of Positive Symptoms; SANS = Scale for the Assessment of Negative Symptoms; CPZ = chlorpromazine-equivalent dose.

### MRI acquisition and processing

T1-weighted images were acquired on two different MRI scanners (GE Medical Systems, Milwaukee, USA) at MR-center, Karolinska Institutet, Stockholm, Sweden: All baseline and 5-year follow-up scans were acquired on the same GE 1.5T Signa HDxt scanner. Thereafter, all 13-year follow-up scans were acquired on a GE 3T Discovery MR750 scanner. Acquisition parameters are provided in the Supplementary Materials, section 1. T1-weighted images were processed with the longitudinal stream in FreeSurfer v7.4.1 (http://surfer.nmr.mgh.harvard.edu)^44–46^ which performs surface reconstruction of the white/gray and pial surfaces, and creates a within-subject average for robust estimation of longitudinal change. Visual quality control was conducted for within-subject average T1-weighted images and surfaces. In the case of gross artifacts or errors in surface reconstruction, single time points were additionally inspected, and participants were excluded from subsequent analyses (n = 17).

For each longitudinally processed time point, LGI was calculated as the ratio of the area of the pial surface relative to the area of a smooth three-dimensional outer surface as implemented in FreeSurfer. Regions with high LGI can be interpreted as having a high degree of “buried cortex”, such as large gyri and major fissures.^4^

### Statistical analyses

Statistical analyses were conducted in MATLAB v2023a^47^ For all cortex-wide LGI analyses, we controlled the False Discovery Rate (FDR) with the Benjamini-Hochberg method and applied a minimum cluster extent threshold of 50 mm^2^. LGI surface analyses were conducted in fsaverage space across the 149,955 vertices of the cortex label (i.e., excluding the medial wall).

### Demographic and clinical variables

Baseline differences between patients and healthy controls in age, sex, handedness, years of schooling, and IQ were assessed with independent sample t-test and Chi-square tests. Baseline predictors of attrition were examined by comparing attendants and dropouts, stratified by follow-up time point and diagnostic group, with independent sample t-test and Chi-square tests. For patients, baseline differences in clinical variables (i.e., SAPS, SANS, CPZ, duration of illness, age at onset, and GAF) between attendants and dropouts were additionally analyzed per follow-up time point.

### Main longitudinal analysis

A spatio-temporal linear mixed-effects model (ST-LME) was employed to assess differences in longitudinal LGI trajectories between SSD and healthy controls, as implemented in FreeSurfer.^48^ Rather than fitting independent models for each vertex, this approach leverages the spatio-temporal covariance of the data, thereby boosting statistical power. Importantly, it accommodates unbalanced longitudinal designs with missing time points, enabling inclusion of participants with missing data for some time points in the same model. The final statistical model used for the main analyses was specified as follows:

> Main model: LGI_ij_ ∼ Intercept_i_ + Time_ij_ + Group_i_ + Time_ij_-by-Group_i_ + BaselineAge_i_ + Sex_i_

where intercept was modeled as a random effect and the indices *i* and *j* correspond to participants (N = 202) and time points (baseline, 5-, or 13-year follow-up), respectively. Time was defined as years since baseline for each time point (i.e., 0 for baseline, approximately 5 for the 5-year follow-up, and approximately 13 for the 13-year follow-up). The main effect of interest was the time-by-group interaction term, representing the estimated difference in longitudinal LGI change between SSD and healthy controls.^48–50^ From this main model, we report the effects of time (i.e., longitudinal change), group (i.e., differences between SSD and healthy controls at baseline), and baseline age (i.e., associations between age at baseline and LGI). Since scanner and time point were highly collinear (all 13-year follow-up scans were acquired on the second scanner), we did not include scanner as a covariate in the main model. Potential confounding by scanner was addressed in separate sensitivity analyses. In the main analysis, the model was fitted to the complete dataset with all available time points.

### Model fit

To evaluate the ST-LME model fit with respect to the specification of random effects, we further fitted a model with two random effects for intercept and time.^48–50^ This model failed to converge in > 40% of regions. Furthermore, we assessed the influence of adding nonlinear effects of age and longitudinal time. Two additional models were fitted, one including baseline age^2^ and one including time^2^ and group x time^2^. There was no significant group x time^2^ interaction. Similarly, age^2^ was significant in only one small cluster. Thus, to ensure a parsimonious model and enhance interpretability, the nonlinear baseline age and time terms were excluded from subsequent analyses.

### Sensitivity analyses

To test the robustness of the findings, we performed a comprehensive set of sensitivity analyses. To assess the impact of the scanner change for the 13-year follow-up, we employed two different approaches. First, we repeated the main analysis where we included scanner as a covariate. Second, we fitted the model to cross-sectionally processed data collected on the same scanner (i.e., baseline and 5-year follow-up data only). To examine the effect of surface reconstruction quality, we repeated the main analysis after excluding 14 participants with high Euler numbers (i.e., a measure of topological surface artifacts reflecting low surface reconstruction quality) determined by outlier detection with a median absolute deviation (MAD) threshold of 4. To assess the influence of the longitudinal processing pipeline, we repeated the main analysis with data from the cross-sectional pipeline of FreeSurfer. To further rule out toolbox-specific effects, we performed surface reconstruction with FastSurfer, which employs a different algorithm for surface reconstruction^51^ and repeated the main analysis. To assess the robustness of the statistical modelling approach, we performed the main analysis with a conventional vertex-wise LME (V-LME) model instead of the ST-LME model.^49^ Detailed results from the sensitivity analyses are available in the Supplementary Materials (Figure S2 – S7).

### Associations with medication and symptom severity

In exploratory analyses, we investigated the relationship between LGI and symptom severity (i.e., total SAPS and SANS scores) and CPZ among patients. For these analyses, the time-varying variables were split into a subject-wise mean and a mean-centered variable:^52^ The subject-wise mean was calculated per participant as the within-subject mean of the variable across the available time points, e.g., mean CPZ_i_ = (CPZ_i,TP1_ + CPZ_i,TP2_ + CPZ_i,TP3_)/3 for CPZ for the *i-*th participant (assumed here to have data for three time points). The mean-centered variable was computed for each individual time point by subtracting the mean from the observed value at that time point, e.g., CPZ_i,TP1_ = CPZ_i,TP1_ - mean CPZ_i_, for the *i*-th participant. In the final model, the mean CPZ_i_, captures between-subject differences and therefore reflects cross-sectional effects. Conversely, the mean-centered variable, CPZ_i,j_, captures within-subject variations and therefore reflects longitudinal effects. The complete statistical model for CPZ (and analogously for symptom severity) was thus specified as follows:

> LGI_ij_ ∼ Intercept_i_ + Time_ij_ + Mean CPZ_i_ + Mean-centered CPZ_ij_ + BaselineAge_i_ + Sex_i_

Here, the intercept was modeled as a random effect and the indices *i* and *j* correspond to patients and time points, respectively. The analyses probing associations with clinical variables were performed on data processed with the cross-sectional pipeline due to missing data for some time points.

## Results

### Demographic and clinical data

Out of 231 participants with MRI data, we excluded six due to missing demographical data, six due to MRI processing errors, and 17 due to poor image quality or surface reconstruction errors. The final study sample included 103 patients with SSD and 99 healthy controls at baseline, 50 patients and 57 healthy controls at the 5-year follow-up, and 42 patients and 60 healthy controls at 13-year follow-up. MRI data for all three time points were available for 32 patients and 42 healthy controls. For 43 patients and 24 controls, only baseline MRI data were available. Baseline and 5-year follow-up data (i.e., no 13-year follow-up data) were available from 18 patients and 15 controls, and baseline and 13-year follow-up data (i.e., no 5-year follow-up data) from 10 patients and 18 controls. Sample characteristics are provided in Table 1.

At baseline, there was a significant difference in IQ between patients with SSD and healthy controls (*t*(135) = 4.85, *p* < .001). There was no significant group difference for age, sex, handedness, and years of schooling. Baseline characteristics associated with attrition at follow-up time points were analyzed stratified by group and time point. In patients, there was no significant difference in baseline variables between those returning for the 5-year follow-up and dropouts. Patients attending the 13-year follow-up were significantly younger (mean age at baseline = 38.32, SD = 8.12) than those who did not attend the 13-year follow-up (mean age at baseline = 43.32, SD = 6.7; *t*(101) = -3.41, *p* < .001). There was no significant difference in any of the remaining variables. Among healthy controls, there was a significant difference in the proportion of right-handed participants in the 5-year follow-up (attendants 87.5% right-handed, dropouts 100%, *X^2^*(1) = 5.52, *p* = .019) and the 13-year follow-up (attendants 88.1% right-handed, dropouts 100%, *X^2^*(1) = 4.86, *p* = .028).

### Main longitudinal analysis

The main analysis revealed no significant differences in longitudinal LGI change between patients with SSD and healthy controls after FDR correction. However, there were significant group differences for the intercept, indicating significantly lower cross-sectional LGI in patients with SSD in widespread frontal, insular, cingulate, and occipital regions across both hemispheres. We observed two restricted clusters of increased cross-sectional LGI in patients relative to controls in the left precuneus and superior parietal cortex (Figure 1 and Table 2). Baseline age had a negative association with LGI, indicating lower LGI with higher age in extensive bilateral clusters in frontal, parietal, and occipital areas (Supplementary Figure S1 & Table S2). Longitudinal change over time revealed a mixed pattern of increases and decreases in LGI. Longitudinal reductions in LGI were apparent in bilateral parietal and cingulate regions, and left insula for both patients and controls. However, substantial longitudinal increases in LGI over time were also observed in the bilateral frontal and left occipital cortex (Supplementary Figure S1 & Table S1).

**Figure 1.**
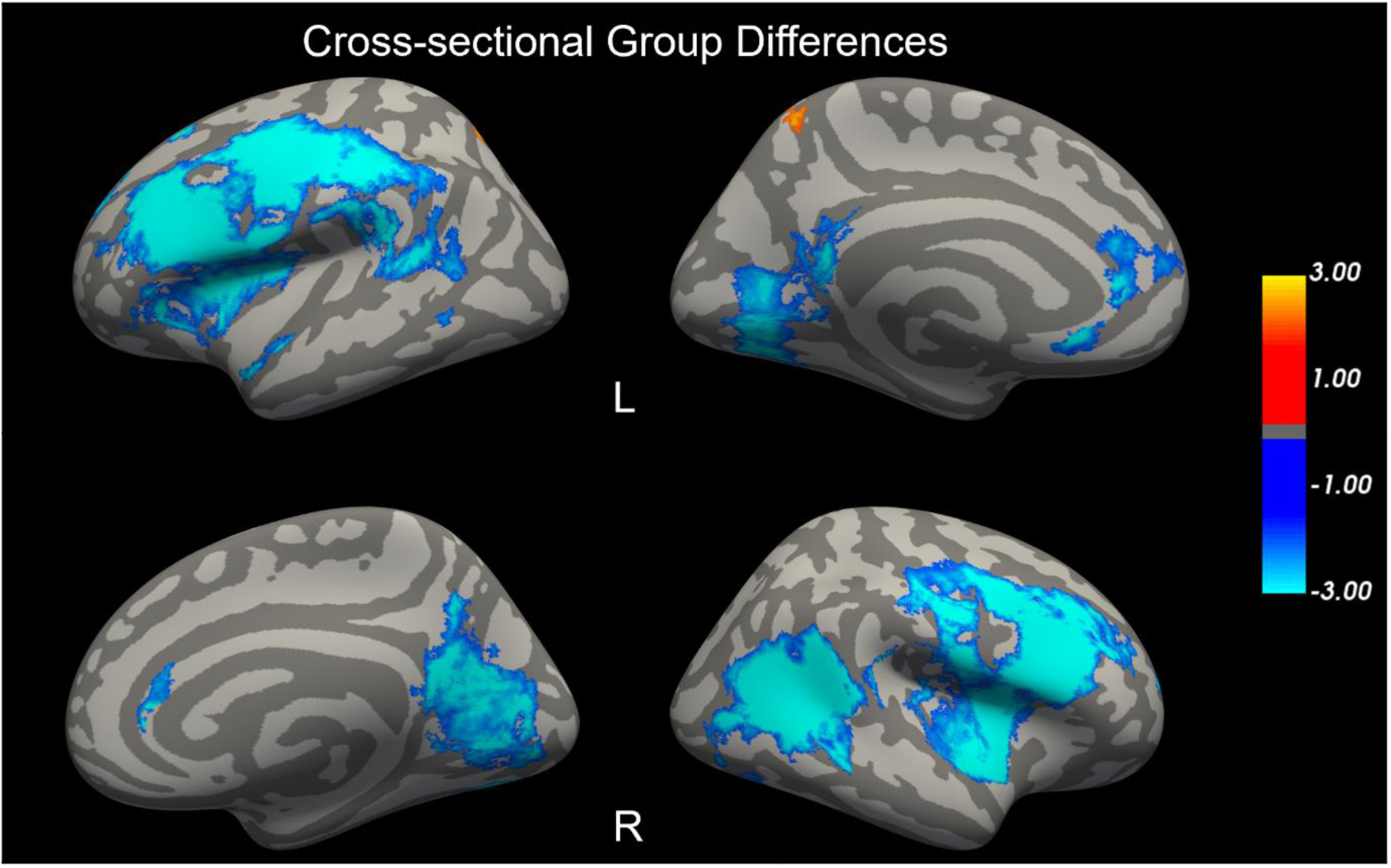
Cross-sectional group differences in Local Gyrification Index (LGI) between patients with schizophrenia spectrum disorder and healthy controls at baseline. Negative log10(p-values) after FDR-correction with p < .05 and minimum cluster extent threshold = 50 mm^2^ for the left (L) and right (R) hemispheres. Blue = patients < controls, red = patients > controls.

**Table 2.** Clusters with significant cross-sectional group differences between patients with schizophrenia spectrum disorder and healthy controls.

| Peak Annotation | Size (mm<sup>2</sup>) | Signed<br>-log<sub>10</sub>(p-value) | Peak Talairach Coordinates |  |  |
| --- | --- | --- | --- | --- | --- |
|  |  |  | X | Y | Z |
| L Rostral middle frontal | 11906 | -7.2 | -40 | 28 | 30 |
| L Lingual | 2423 | -4.1 | -22 | -74 | -1 |
| L Rostral anterior cingulate | 457 | -4.56 | -5 | 33 | 1 |
| L Rostral middle frontal | 215 | -3.87 | -21 | 48 | 27 |
| L Superior temporal | 214 | -3.43 | -57 | 0 | -7 |
| L Superior frontal | 151 | -5.02 | -23 | 27 | 47 |
| L Rostral anterior cingulate | 122 | -3.46 | -6 | 23 | -6 |
| L Precuneus | 109 | 2.73 | -9 | -51 | 54 |
| L Superior parietal | 59 | 2.81 | -16 | -61 | 48 |
| L Middle temporal | 51 | -2.74 | -58 | -53 | 1 |
| R Insula | 8642 | -5.8 | 34 | 6 | 12 |
| R Inferior parietal | 3559 | -5.79 | 53 | -52 | 11 |
| R Precuneus | 3545 | -4.24 | 7 | -52 | 18 |
| R Fusiform | 248 | -4.75 | 30 | -72 | -5 |
| R Supramarginal | 143 | -3.05 | 44 | -33 | 25 |
| R Fusiform | 138 | -2.96 | 41 | -67 | -9 |
| R Rostral anterior cingulate | 109 | -3.39 | 8 | 33 | 2 |
| R Rostral middle frontal | 79 | -3.72 | 23 | 55 | 9 |
| R Lateral occipital | 52 | -2.98 | 44 | -76 | -6 |

*Note.* L = left hemisphere, R = right hemisphere.

### Sensitivity analyses

When including scanner as a covariate in the model, we similarly observed no differences in longitudinal change between SSD and healthy controls. The cross-sectional group difference aligned with that of the main analysis and showed lower LGI in patients in bilateral frontal, occipital and insular regions. The model indicated a strong association between scanner and LGI, with higher LGI in the right frontal cortex for the second scanner. In contrast to the main analysis, we observed no significant longitudinal change over time (i.e., no effect for time) when adjusting for scanner. Baseline age was again associated with decreased LGI in large areas covering frontal, parietal, and occipital regions (Supplementary Figure S2).

The ST-LME model with cross-sectionally processed data acquired on the same scanner (i.e., baseline and 5-year follow-up only) likewise showed no significant differences in longitudinal change between patients with SSD and healthy controls. In line with the main analysis, we found reduced cross-sectional LGI in SSD compared to healthy controls in similar widespread regions. In contrast to the main analysis, there was no significant longitudinal effect of time. Baseline age showed similar negative cross-sectional associations with LGI in extensive frontal, parietal, and occipital clusters (Supplementary Figure S3).

We observed only minor differences in the sensitivity analyses after excluding Euler number outliers, when using data from the cross-sectional FreeSurfer pipeline, statistical modeling with the V-LME model instead of the ST-LME model or using LGI data processed with FastSurfer. Importantly, there were no significant differences in longitudinal change in LGI between SSD and healthy controls, thus corroborating the findings of the main analysis. These results are reported in the Supplementary Materials (Figure S4 – S7).

### Associations with medication and symptom severity

In the patient sample, we found a negative cross-sectional association with mean CPZ in the left medial orbitofrontal cortex (OFC), extending into the left rostral anterior cingulate cortex (rACC), and in a cluster in the left postcentral gyrus with cross-sectionally processed data (Figure 2 and Table 3). We observed neither longitudinal associations with mean-centered CPZ nor cross-sectional or longitudinal associations with positive or negative psychotic symptom severity.

**Figure 2.**
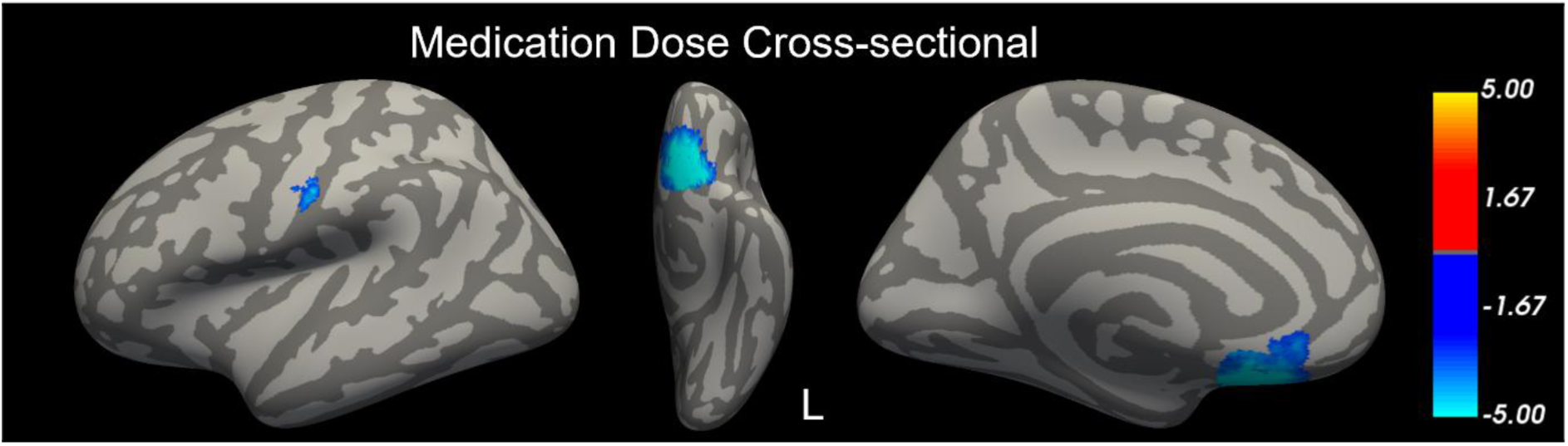
Association between the Local Gyrification Index (LGI) and chlorpromazine-equivalent medication dose (CPZ) among patients with schizophrenia spectrum disorders. Negative log10(p-values) after FDR-correction with p < .05 and minimum cluster extent threshold = 50 mm^2^ for the left hemisphere (L).

**Table 3.** Clusters of cross-sectional associations with chlorpromazine-equivalent medication dose among patients with schizophrenia spectrum disorders.

| Peak Annotation | Size (mm<sup>2</sup>) | Signed<br>-log<sub>10</sub>(p-value) | Peak Talairach Coordinates |  |  |
| --- | --- | --- | --- | --- | --- |
|  |  |  | X | Y | Z |
| L Rostral anterior cingulate | 1101 | -8.88 | -5 | 14 | -6 |
| L Postcentral | 137 | -4.74 | -59 | -9 | 32 |

*Note.* L = Left hemisphere.

## Discussion

In this 13-year longitudinal study, patients with chronic SSD showed no deviation from the age-related changes in gyrification trajectories observed in healthy controls. This finding was robust with respect to a variety of MRI processing and statistical modeling approaches. In line with previous studies, we observed extensive cross-sectional hypogyria in SSD relative to controls at baseline. Higher antipsychotic medication dose was not associated with longitudinal change in LGI but was associated cross-sectionally with lower LGI among patients. We observed no significant associations between LGI and psychotic symptom severity.

Our main finding that cortical folding trajectories do not significantly diverge in patients with SSD compared to healthy controls contradicts models that predict an accelerated age-related decline in chronic SSD.^13,15^ However, this hypothesis is primarily based on cross-sectional studies reporting higher cortical folding in early illness stages and lower cortical folding in chronic patients relative to controls. In contrast, a recent large-scale study of cortical morphology in adolescents with early-onset psychosis reported an extensive pattern of lower LGI in patients compared to healthy adolescents.^53^ Moreover, further recent studies did not observe the proposed hypergyria in individuals with high familial risk^54^ and first-episode psychosis.^55^ These findings suggest that earlier reports of hypergyria in early illness stages may not generalize across all populations and warrant further investigation.

Previous longitudinal studies in adolescent-onset psychosis^20^ and in first-episode psychosis^19^ with shorter follow-up durations (up to 2.7 years) have reported a steeper decline in patients compared to healthy controls. Although we did not observe such progressive alterations of LGI in chronic SSD, we cannot rule out its occurrence in individuals at risk or in earlier stages of the illness. Another possibility is that lower LGI reflects neurodevelopmental disruptions present before illness onset. This would be in line with the two-hit hypothesis, whereby early disturbances of brain development (first hit)^56^, e.g., manifesting as aberrant gyrification, later interact with maturational and environmental stressors (second hit) to trigger the onset of a psychotic disorder. As such, our results are consistent with the interpretation of cortical folding as a marker of neurodevelopmental disruptions.^5,6^ The discrepancy between observed hypergyria in individuals at risk and hypogyria in chronic SSD should be addressed in future long-term follow-up studies. Further longitudinal studies in risk populations, and early illness stages are needed to disentangle the contribution of dynamic relative to pre-existing static cortical folding alterations.

In line with cross-sectional studies in chronic SSD, we observed extensive cross-sectional patterns of lower LGI in patients compared to healthy controls at baseline. Lower LGI was seen across widespread areas in the bilateral frontal, occipital, and insular cortices. These results are in agreement with a previous cross-sectional study employing a partly overlapping sample, that additionally showed hypogyria in a second sample with a shorter duration of illness of 4.7 years.^18^ Unlike this previous study, we employed both baseline and follow-up data and a different statistical approach designed to enhance statistical power in vertex-wise analyses. Thus, while the spatial concordance of the cross-sectional LGI deficits was high, the regional extent of the effects was slightly greater in the present study. The findings further align with those of other studies in chronic SSD that have also reported hypogyria.^57–63^ This agreement with prior cross-sectional findings supports the methodological robustness of the present study and our findings provide further evidence of hypogyria in chronic SSD. The present study extends these findings by indicating that these LGI deficits in patients with chronic SSD are not exacerbated over time.

We observed cross-sectional associations among patients indicating that higher CPZ was associated with lower LGI in the left OFC, extending into the rACC, with a separate cluster in the left postcentral gyrus. A prior study in first-episode psychosis reported similar negative association with antipsychotic medication use, albeit across broader regions. Notably, they reported associations between LGI in the left OFC and rACC for antipsychotic medication duration but not dose.^24^ Conversely, another study in minimally medicated patients reported an association with increased LGI in right OFC after 1-11 months of medication,^17^ although the duration of medication exposure in this study was shorter than in the current sample. Several studies have reported no association between cortical folding and antipsychotic medication.^26,27,57,61^ This includes the previous cross-sectional assessment on the current sample, although the association to medication was only assessed in clusters with significant group differences.^18^ Given the well-established impact of antipsychotic medication on other structural brain measures,^64,65^ further research is needed to unravel the inconsistent findings for gyrification.

An important limitation of the study was the MRI scanner change at the 13-year follow-up. Indeed, the positive longitudinal effects in frontal and superior areas (Supplementary Figure S1 and Table S1) were absent when analyses were limited to data from the same scanner, suggesting that they reflected scanner-related differences. This interpretation aligns with prior studies showing longitudinal declines of gyrification over time.^10–12^ While the effects of time and scanner are not easily disentangled, the primary result of no accelerated decline in SSD was replicated in same-scanner data (i.e., baseline and 5-year follow-up). Thus, it is unlikely that group differences in scanner effects explained these results. MRI studies have increasingly employed scanner harmonization procedures to adjust for scanner-related differences. However, since all 13-year data were acquired on the second scanner, time and scanner effects are not statistically separable, precluding the use of existing scanner harmonization methods.^66^ Nonetheless, sensitivity analyses including scanner as a covariate replicated the primary result. Scanner upgrades or changes are common in long-term follow-up studies and future research is needed to develop new methods to alleviate the influence of scanner-related differences.

Additional limitations include attrition over the extended follow-up period. Younger patients were more likely to return at the 13-year follow-up, narrowing the available age range for longitudinal analyses. Survivorship bias may also have influenced the findings, as more severely ill patients may have been less likely to participate at later time points. Moreover, information on antipsychotic medication use and psychotic symptom trajectories between assessments was limited. In addition, the sex distribution of the sample was unbalanced. While we corrected for sex in the analyses, the modest number of female participants did not allow us to assess sex differences for longitudinal trajectories. Furthermore, the sample consisted of individuals with chronic schizophrenia (mean duration of illness at baseline: 16.37 years), precluding the estimation of gyrification changes occurring prior to or shortly after illness onset. Future research should aim to characterize putative progressive gyrification changes in psychotic disorders in developmental samples or with earlier onset and address potential sex differences.

## Conclusions

Based on a large sample with a uniquely long follow-up period of over 13 years, we found that while cortical folding is reduced in individuals with chronic SSD, there was no evidence of progressive decline beyond that expected from normal aging. These results challenge the prevailing hypothesis of dynamic cortical folding changes across the lifespan in chronic SSD.^13^ Instead, our findings support the interpretation of reduced gyrification as a static marker of early neurodevelopmental disruption, rather than a dynamic indicator of ongoing neurodegeneration. These results align with emerging evidence that lifelong brain structural variation may be primarily shaped by genetic liability and early-life developmental factors, e.g., birth weight, rather than by degenerative processes occurring later in life.^67^ Taken together, our findings reinforce the interpretation of hypogyria in SSD as a neurodevelopmental phenotype, with implications for the conceptualization of structural imaging markers in neuroimaging research on SSD.

## Supporting information

Supplement_Wallenwein_Longitudinal_LGI_ChronicSSD

## Data Availability

The data supporting the findings of this study may be obtained from the corresponding author upon reasonable request.

## Acknowledgements

We are grateful to the participants in the study, as well as the clinicians in the HUBIN project involved in recruitment and clinical assessments. Data processing and analysis was conducted on the TSD (Tjenester for Sensitive Data) facilities, owned by the University of Oslo, operated and developed by the TSD service group at the IT department (USIT) of the University of Oslo.

## Author contributions

**L.A.Wa.**: Conceptualization, Formal Analysis, Methodology, Software, Visualization, Writing – Original Draft, Writing – Review & Editing; **L.A.Wo.**: Writing – Review & Editing; **C.B.**: Data Curation, Writing – Review & Editing; **P.P.**: Writing – Review & Editing; **K.N.J.**: Writing – Review & Editing; **L.P.**: Writing – Review & Editing; **D.M.**: Writing – Review & Editing, Supervision; **E.G.J.**: Investigation, Resources, Data Curation, Writing – Review & Editing, Funding Acquisition; **I.A.**: Resources, Project Administration, Funding Acquisition, Writing – Review & Editing, Supervision; **S.N.**: Conceptualization, Data Curation, Methodology, Project Administration, Writing – Review & Editing, Supervision.

