## Supplement_Wallenwein_Longitudinal_LGI_ChronicSSD for "Longitudinal trajectories of cortical folding in schizophrenia spectrum disorders: a 13-year follow-up study"

### 1. Imaging parameters

Baseline and 5-year follow-up T1-weighted images were acquired on a General Electric 1.5-Tesla Signa HDxt scanner with an 8-channel head coil and a three-dimensional spoiled gradient recalled (SPGR) pulse sequence. Imaging parameters were as follows: 124 coronal slices, 35° flip angle, repetition time 24 ms, echo time 6.0 ms, voxel size 0.86 × 0.86 × 1.50 mm, acquisition matrix = 256 × 192, field of view = 24 cm. Images for the 13-year follow up were acquired on a General Electric 3-Tesla Discovery MR750 scanner and a 32-channel head coil. T1-weighted images were acquired with a three-dimensional gradient echo sequence and the following parameters: sagittal orientation, flip angle = 12°, repetition time = 7.90 ms, echo time = 3.1 ms, inversion time = 450 ms, slice thickness = 1.2 mm, bandwidth = 244.14 Hz/pixel, voxel size = 1 × 1 × 1.2 mm.

### 2. Slope and effect of baseline age in ST-LME model with longitudinal processing

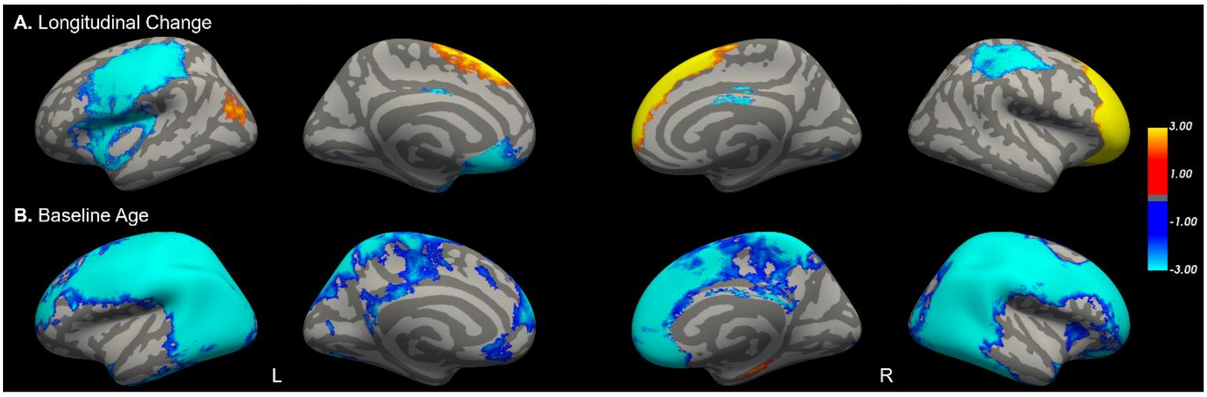


**Figure S1.** Significant effects in the spatio-temporal linear mixed effects model based on longitudinally processed surfaces. **A.** Longitudinal change over time. **B.** Effect of baseline age. Negative log10(p-values) after FDR-correction with $p < .05$ and minimum cluster extent threshold = 50 mm$^2$ for the left (L) and right (R) hemispheres.

**Table S1.** Clusters with a significant effect of longitudinal change over time.

| Peak Annotation | Size ($mm^2$) | Signed -log10(p-value) | Peak Talairach Coordinates | | |
|---|---|---|---|---|---|
| | | | X | Y | Z |
| L Postcentral | 12806 | -12.61 | -44 | -18 | 49 |
| L Rostral anterior cingulate | 2023 | -10.11 | -5 | 27 | -4 |
| L Superior frontal | 1639 | 6.53 | -10 | 13 | 45 |
| L Inferior parietal | 636 | 3.67 | -37 | -83 | 17 |
| L Entorhinal | 456 | -7.12 | -28 | 0 | -24 |
| L Fusiform | 107 | -2.72 | -40 | -28 | -17 |
| L Lateral occipital | 70 | 2.17 | -18 | -98 | 7 |
| L Posterior cingulate | 55 | -6.12 | -5 | 6 | 29 |
| R Rostral middle frontal | 10912 | 30 | 34 | 49 | 5 |
| R Precentral | 4144 | -8.12 | 41 | -3 | 50 |
| R Caudal anterior cingulate | 115 | -9.17 | 3 | 1 | 27 |
| R Lingual | 61 | -2.73 | 13 | -78 | -6 |
| R Posterior cingulate | 52 | -4.65 | 4 | -11 | 34 |

*Note.* L = left hemisphere, R = right hemisphere.

**Table S2.** Clusters with a significant effect of baseline age.

| Peak Annotation | Size ($mm^2$) | Signed -log10(p-value) | Peak Talairach Coordinates | | |
|---|---|---|---|---|---|
| | | | X | Y | Z |
| L Supramarginal | 42119 | -30 | -49 | -45 | 43 |
| L Rostral anterior cingulate | 407 | -3.24 | -5 | 25 | -4 |
| L Superior temporal | 259 | -4.15 | -39 | 9 | -25 |
| L Superior frontal | 133 | -2.24 | -10 | 23 | 35 |
| L Pericalcarine | 100 | -2.9 | -15 | -78 | 12 |
| L Medial orbitofrontal | 62 | -1.93 | -10 | 33 | -19 |
| L Rostral middle frontal | 59 | -2.47 | -41 | 37 | 1 |
| L Lateral orbitofrontal | 57 | -3.66 | -27 | 9 | -15 |
| R Supramarginal | 46395 | -30 | 50 | -29 | 40 |
| R Superior temporal | 498 | -5.45 | 38 | 2 | -22 |
| R Insula | 392 | -2.52 | 32 | 19 | 1 |
| R Entorhinal | 190 | 4.56 | 23 | -8 | -27 |
| R Posterior cingulate | 83 | -4.96 | 6 | -10 | 29 |

*Note.* L = left hemisphere, R = right hemisphere.

### 3. Sensitivity analyses

#### 3.1. ST-LME longitudinal processing with scanner as covariate

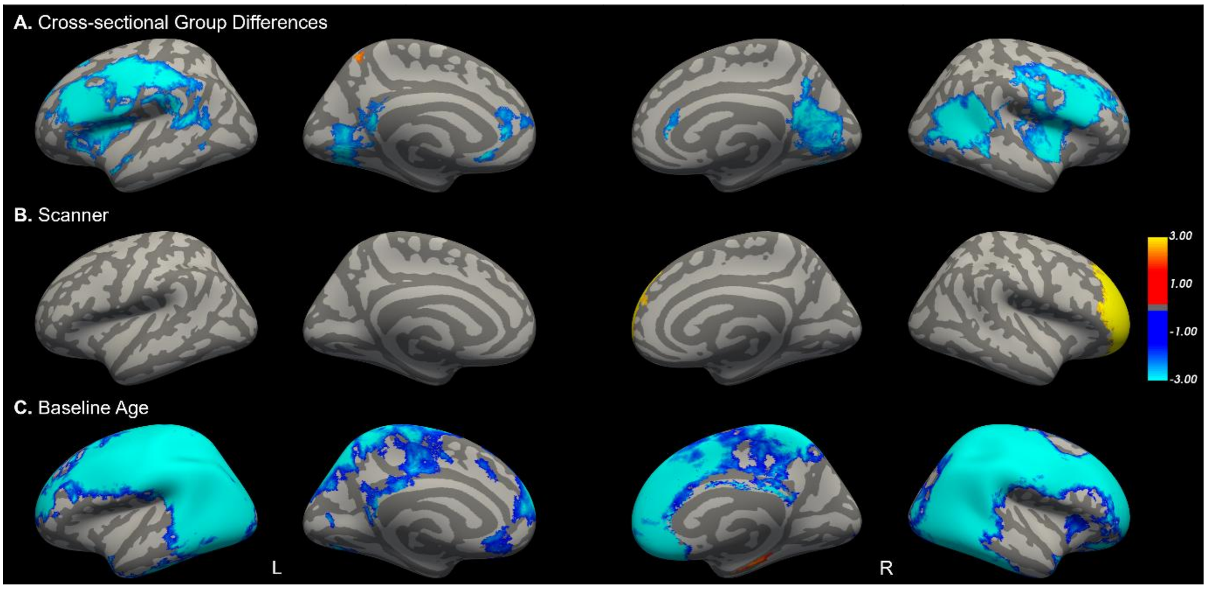


**Figure S2.** Significant effects in the spatio-temporal linear mixed effects model based on longitudinally processed surfaces including scanner as a covariate. No significant longitudinal change or time-by-group interaction was observed. **A.** Cross-sectional group differences between patients with schizophrenia spectrum disorder and healthy controls at baseline. **B.** Effect of scanner. **C.** Effect of baseline age. Negative log10(p-values) after FDR-correction with $p < .05$ and minimum cluster extent threshold = 50 $mm^2$ for the left (L) and right (R) hemispheres.

#### 3.2. ST-LME cross-sectional processing without 13-year follow-up

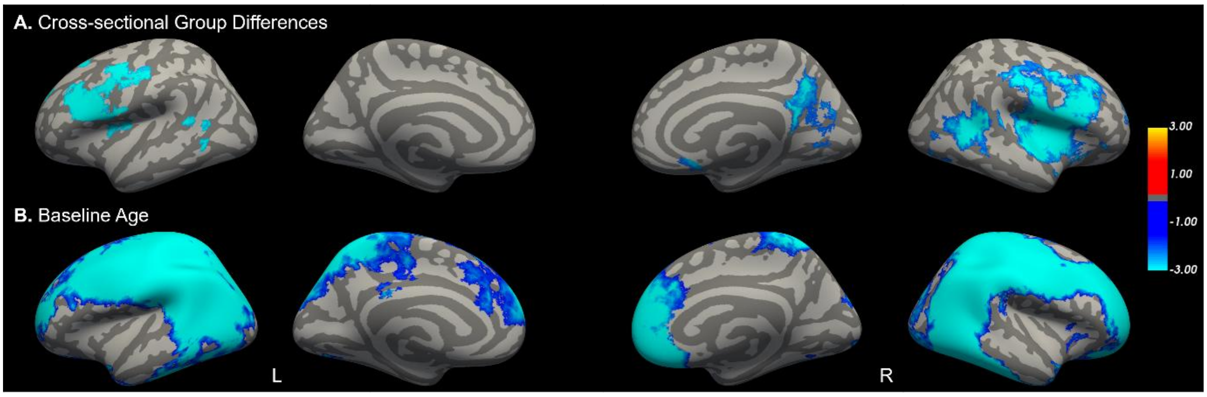


**Figure S3.** Significant effects in the spatio-temporal linear mixed effects model based on cross-sectionally processed surfaces with data of the baseline and 5-year follow-up only. No significant longitudinal change or time-by-group interaction was observed. **A.** Cross-sectional group differences between patients with schizophrenia spectrum disorder and healthy controls at baseline. **B.** Effect of baseline age. Negative log10(p-values) after FDR-correction with $p < .05$ and minimum cluster extent threshold = 50 $mm^2$ for the left (L) and right (R) hemispheres.

#### 3.3. ST-LME longitudinal processing excluding outliers in Euler number

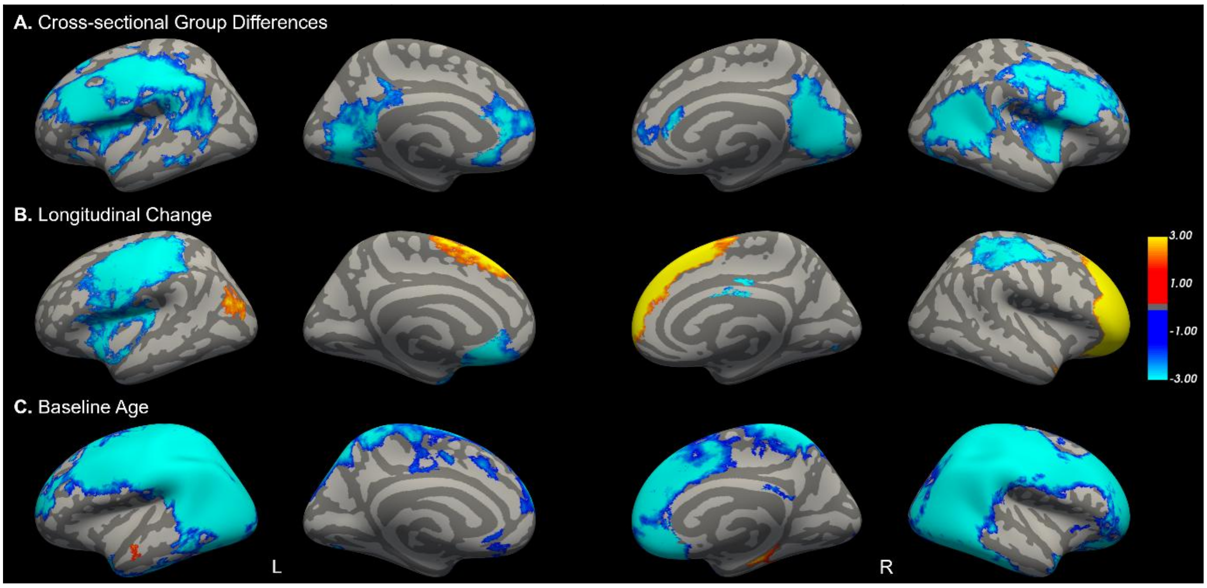


**Figure S4.** Significant effects in the spatio-temporal linear mixed effects model based on longitudinally processed surfaces after exclusions of outliers in Euler number. No significant time-by-group interaction was observed. **A.** Cross-sectional group differences between patients with schizophrenia spectrum disorder and healthy controls at baseline. **B.** Longitudinal change over time. **C.** Effect of baseline age. Negative log10(p-values) after FDR-correction with $p < .05$ and minimum cluster extent threshold = 50 $mm^2$ for the left (L) and right (R) hemispheres.

#### 3.4. ST-LME cross-sectional processing

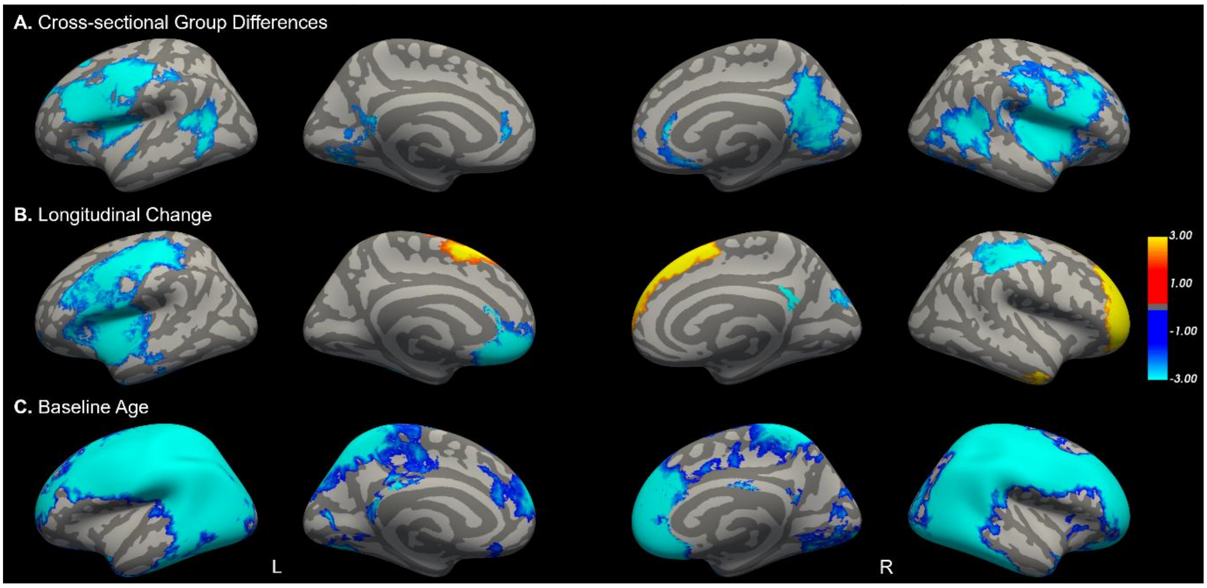


**Figure S5.** Significant effects in the spatio-temporal linear mixed effects model based on cross-sectionally processed surfaces. No significant time-by-group interaction was observed. **A.** Cross-sectional group differences between patients with schizophrenia spectrum disorder and healthy controls at baseline. **B.** Longitudinal change over time. **C**. Effect of baseline age. Negative log10(p-values) after FDR-correction with $p < .05$ and minimum cluster extent threshold = 50 $mm^2$ for the left (L) and right (R) hemispheres.

#### 3.5. ST-LME FastSurfer cross-sectional processing

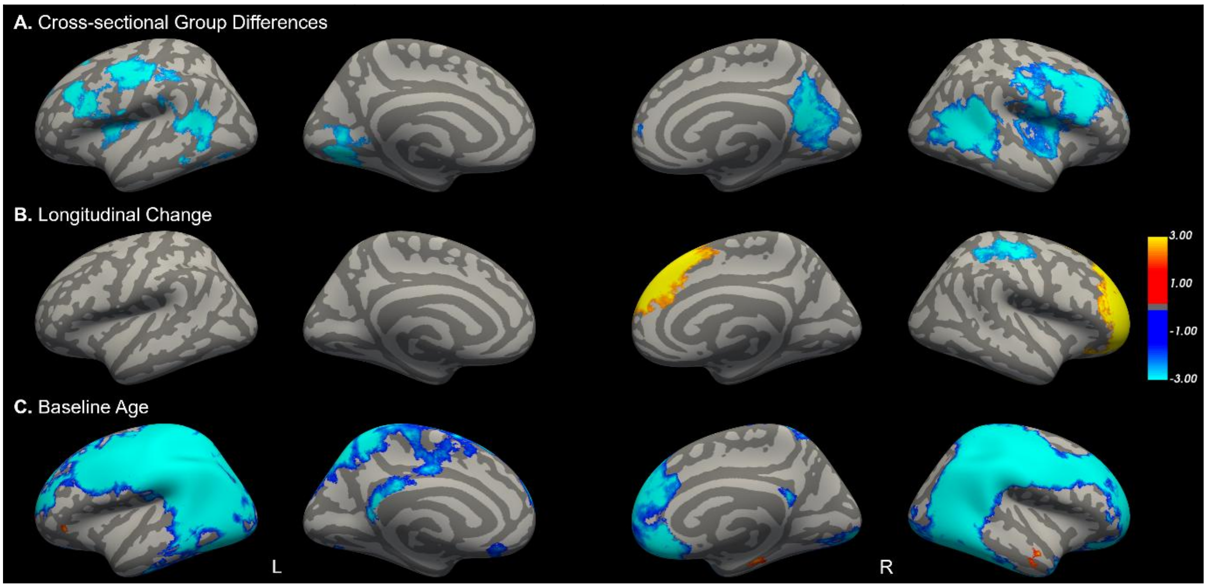


**Figure S6.** Significant effects in the spatio-temporal linear mixed effects model based on cross-sectionally processed surfaces with FastSurfer. Four participants were excluded for this analysis due to processing errors. No significant time-by-group interaction was observed. **A.** Cross-sectional group differences between patients with schizophrenia spectrum disorder and healthy controls at baseline. **B.** Longitudinal change over time. **C.** Effect of baseline age. Negative log10(p-values) after FDR-correction with $p < .05$ and minimum cluster extent threshold = 50 $mm^2$ for the left (L) and right (R) hemispheres.

#### 3.6. V-LME longitudinal processing

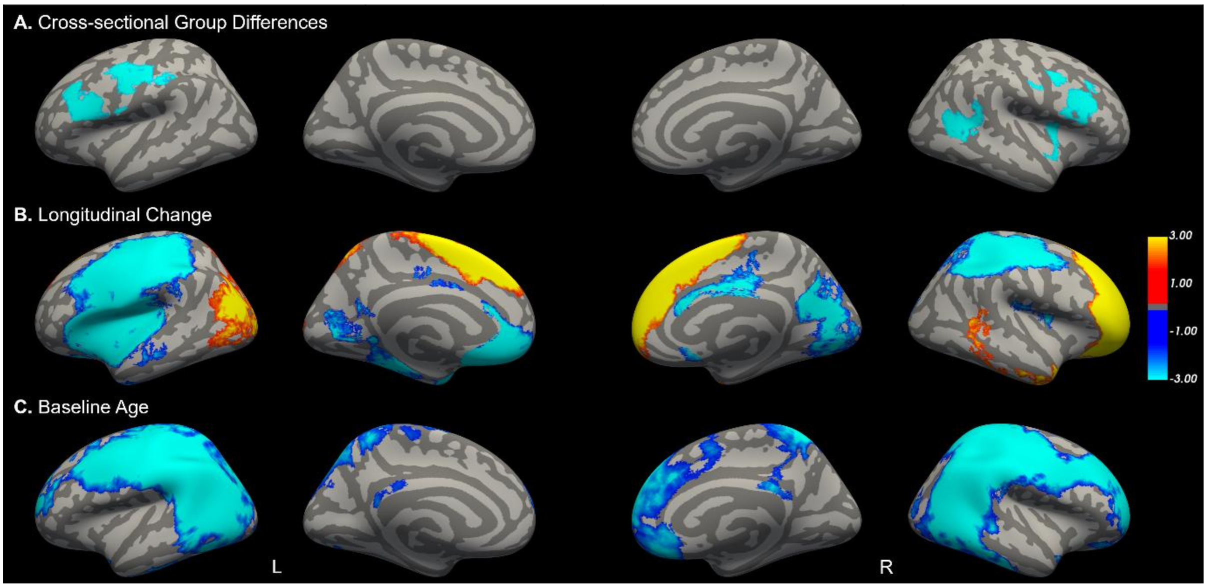


**Figure S7.** Significant effects in the vertex-wise linear mixed effects model based on longitudinally processed surfaces. No significant time-by-group interaction was observed. **A.** Cross-sectional group differences between patients with schizophrenia spectrum disorder and healthy controls at baseline. **B.** Longitudinal change over time. **C.** Effect of baseline age. Negative log10(p-values) after FDR-correction with $p < .05$ and minimum cluster extent threshold = 50 $mm^2$ for the left (L) and right (R) hemispheres.
